# An exploratory randomized controlled study on the efficacy and safety of lopinavir/ritonavir or arbidol treating adult patients hospitalized with mild/moderate COVID-19 (ELACOI)

**DOI:** 10.1101/2020.03.19.20038984

**Authors:** Yueping Li, Zhiwei Xie, Weiyin Lin, Weiping Cai, Chunyan Wen, Yujuan Guan, Xiaoneng Mo, Jian Wang, Yaping Wang, Ping Peng, Xudan Chen, Wenxin Hong, Guangming Xiao, Jinxin Liu, Lieguang Zhang, Fengyu Hu, Feng Li, Fuchun Zhang, Xilong Deng, Linghua Li

**Author notes:** Correspondence to: Linghua Li, Infectious Disease Center, Guangzhou Eighth People’s Hospital, Guangzhou Medical University, Guangzhou, 510060, China. or Xilong Deng, Intensive Care Unit, Guangzhou Eighth People’s Hospital, Guangzhou Medical University, Guangzhou 510440, China. or Fuchun Zhang, Infectious Disease Center, Guangzhou Eighth People’s Hospital, Guangzhou Medical University, Guangzhou, 510060, China. Yueping Li, Zhiwei Xie, Weiyin Lin and Weiping Cai contributed equally.

## Abstract

**Background:** Antiviral therapies against the novel coronavirus SARS-CoV-2, which has caused a global pandemic of respiratory illness called COVID-19, are still lacking.

**Methods:** Our study (NCT04252885, named ELACOI), was an exploratory randomized (2:2:1) controlled trial assessing the efficacy and safety of lopinavir/ritonavir (LPV/r) or arbidol monotherapy for treating patients with mild/moderate COVID-19.

**Findings:** This study successfully enrolled 86 patients with mild/moderate COVID-19 with 34 randomly assigned to receive LPV/r, 35 to arbidol and 17 with no antiviral medication as control. Baseline characteristics of the three groups were comparable. The primary endpoints, the average time of positive-to-negative conversion of SARS-CoV-2 nucleic acid and conversion rates at days 7 and 14, were similar between groups (all *P*>0.05). There were no differences between groups in the secondary endpoints, the rates of antipyresis, cough alleviation, or improvement of chest CT at days 7 or 14 (all *P*>0.05). At day 7, eight (23.5%) patients in the LPV/r group, 3 (8.6%) in the arbidol group and 2(11.8%) in the control group showed a deterioration in clinical status from moderate to severe/critical(*P* =0.206). Overall, 12 (35.3%) patients in the LPV/r group and 5 (14.3%) in the arbidol group experienced adverse events during the follow-up period. No apparent adverse event occurred in the control group.

**Conclusions:** LPV/r or arbidol monotherapy present little benefit for improving the clinical outcome of patients hospitalized with mild/moderate COVID-19 over supportive care.

**Funding:** This study was supported by project 2018ZX10302103-002, 2017ZX10202102-003-004 and Infectious Disease Specialty of Guangzhou High-level Clinical Key Specialty (2019-2021).

## Introduction

Since December 2019, a novel coronavirus disease (COVID-19) has rapidly spread throughout the world, with outbreaks in more than 200 countries and regions [1,2]. The pathogenic organism responsible was identified and termed severe acute respiratory syndrome coronavirus 2 (SARS-CoV-2, previously named 2019 novel coronavirus or 2019-nCoV), belonging to the same family of viruses responsible for the severe acute respiratory syndrome (SARS) and Middle East respiratory syndrome (MERS) [3]. The World Health Organization (WHO) declared COVID-19 as a public health emergency of international concern and characterized the outbreak as a pandemic on March 12, 2020. As of March 31^st^, 2020, 750,890 confirmed cases and 36,405 death cases have been documented globally [2]. Despite its rapid global spread, little is known about the pathogenesis of the virus or its infectious host. Even worse, no vaccine or specific antiviral drugs have demonstrated efficacy in prevention or treatment of COVID-19, which has led to great difficulty in controlling the epidemic and decreasing the mortality rate [4].

Based on “Diagnosis and treatment of pneumonitis caused by new coronavirus (trial version 6)” issued by the National Health Commission of China on February 19^th^ 2020, several drugs, including lopinavir/ritonavir (LPV/r) and arbidol, were recommended as antiviral regimens for the treatment of COVID-19 [5]. Lopinavir is a human immunodeficiency virus 1 (HIV-1) protease inhibitor, usually combined with ritonavir to inhibit cytochrome P450 in order to increase the half-life of lopinavir [6]. In the past 10 years, LPV/r has been proven to have good efficacy and limited side effects for treating HIV-1[7]. Lopinavir was reported to have antiviral activity against MERS-CoV in Vero cells (concentration causing a 50% reduction in replication (EC50) = 8 µM) [8]. Additionally, despite lacking a valid estimate of efficacy, the combination of LPV/r has been associated with significantly fewer adverse clinical outcomes (acute respiratory distress syndrome or death) in 41 patients with SARS compared with ribavirin alone in 111 historical controls (2.4% versus 28.8%, *P*= 0.001) in the 21 days after the onset of symptoms [9]. Thus, based on in vitro testing and previous clinical trials demonstrating its efficacy against other coronaviruses, LPV/r was regarded as an option for treating COVID-19.

Arbidol is a haemagglutinin inhibitor that can effectively block the fusion of influenza virus with its host cell. In addition, it can also induce the immune system to produce endogenous interferon against virus replication, enhance the phagocytic function of macrophages, and activate natural killer cells. [10, 11]. Arbidol was reported to be effective against all strains of influenza viruses (A, B, C), especially influenza A viruses (H1N1, H2N2, H3N3), and to be safe with few side effects [12]. Arbidol was also shown to have direct antiviral effects by inhibiting the replication of SARS virus in vitro [13].

Despite the above preliminary evidence, the actual clinical efficacy of LPV/r or arbidol against SARS-CoV-2 are unknown. Therefore, we were urgently in need of a randomized clinical trial (RCT) to evaluate the efficacy or adverse outcomes of LPV/r or arbidol for treating COVID-19. Guangzhou Eighth People’s Hospital is a designated hospital for the treatment of COVID-19 patients and over 80% of the patients confirmed with COVID-19 in Guangzhou were hospitalized at this facility. Here, we report an exploratory randomized and controlled study (ELACOI) at this hospital, aiming to provide a preliminary evaluation of the efficacy and safety of monotherapy with LPV/r or arbidol in the treatment of patients with mild/moderate COVID-19.

## STAR Methods

### LEAD CONTACT

Further information and requests for resources and reagents should be directed to and will be fulfilled by the Lead Contact, Linghua Li. This study did not generate new unique reagents and did not generate new datasets.

## EXPERIMENTAL MODEL AND SUBJECT DETAILS

### Study design and participants

ELACOI was a single-center, randomized and controlled trial conducted at Guangzhou Eighth People’s Hospital to preliminarily investigate the efficacy of LPV/r and arbidol in treating patients with COVID-19. This empirically exploratory study was initially designed to enroll 125 patients, based on the estimated number of patients admitted to the hospital. We ultimately recruited 86 patients, who were randomly assigned (2:2:1) into 3 groups as follows: In group A (LPV/r group), 34 patients were administered lopinavir (200mg) boosted by ritonavir (50mg) (orally administed, twice daily, 500 mg, each time for 7-14 days). In group B (arbidol group), 35 patients were given arbidol (100mg) (orally administed, 200mg three times daily for 7-14 days). In group C (control group), 17 patients were not given any antiviral therapy. All three groups were followed for up to 21 days. All three groups were treated with supportive care and effective oxygen therapy if in need.^5^ Antiviral treatment was discontinued for patients who 1) had been treated for more than 7 days and tested negative for SARS-CoV-2 nucleic acid in two consecutive tests separated by more than 24 hours, or 2) were discharged from hospital, or 3) had intolerable side effects.

All participants met the following inclusion criteria: 1) age between 18 and 80 years; 2) SARS-CoV-2 infection confirmed by real-time PCR (RT-PCR) from pharyngeal swab; 3) mild clinical status, defined as having mild clinical symptoms but no signs of pneumonia on imaging or moderate clinical status, defined as having fever, respiratory symptoms and pneumonia on imaging [5]; 4) the following lab findings: creatinine ≤110μmol/L, creatinine clearance rate (eGFR) ≥60 ml/min/1.73m^2^, aspartate aminotransferase (AST) and alanine aminotransferase (ALT) ≤5 × ULN, and total bilirubin (TBIL) ≤2 × ULN; 5) willingness to participate in the study and provide informed consent. Patients were excluded based on the following criteria: 1) known or suspected to be allergic to LPV/r or arbidol; 2) having severe nausea, vomiting, diarrhea or other complaints affecting oral intake or absorption in the digestive tract; 3) taking other drugs that may interact with LPV/r or arbidol; 4) having serious underlying diseases, including but not limited to heart, lung, or kidney disease, liver malfunction, or mental illnesses affecting treatment compliance; 5) complications with pancreatitis or hemophilia prior to the trial; 6) Pregnant or lactating women; 7) suspected or confirmed history of alcohol or substance use disorder; 8) participation in other drug trials within the past month; 9) deemed otherwise unsuitable for the study by researchers.

Before initiation of the trial, the protocol was approved by the ethics committee of Guangzhou Eighth People’s Hospital (Approval No. 202002136) and registered on ClinicalTrials.gov (NCT04252885). The ethics committee agreed to set up the control group owing to a lack of reliable evidence about the benefit of present antiviral regimens for treating COVID-19. The trial was also performed in accordance with the International Conference on Harmonization’s Guideline for Good Clinical Practice. Written informed consent was obtained from all screened patients after they fully understood the meaning of the trial and the potential risks involved.

## METHOD DETAILS

### Randomization and masking

All eligible participants were assigned a randomization number which allocated them into one treatment group. The randomization numbers were computer-generated. Allocation concealment was achieved using a centralized web-based randomization system in which the participant identifier (hospitalization number) was entered before the allocation was revealed. The randomization numbers were used in case report form (CRF) pages. The study was blinded to participants and those physicians and radiologists who reviewed the data and radiological images but open-label to clinicians who recruited patients and research staff.

### Procedures

A standardized protocol was developed for collecting clinical data for all participants. The following data were collected: 1) important dates, including fever onset, admission, progression to severe clinical status, positive-to-negative conversion of SARS-CoV-2 nucleic acid, improvement of chest computerized tomography [CT] scan, discharge, or death; 2) presence of predefined comorbidities (hypertension, diabetes mellitus, etc.); 3) daily observation of clinical parameters (temperature, pulse, respiratory rate, oxygen saturation, Inhaled oxygen concentration if needed); 4) The conversion time of nucleic acid of SARS-CoV-2 from positive to negative and the clinical improvement including the rate of antipyresis, the rate of cough alleviation, the rate of improvement on chest CT at days 7 and 14; 5) details of drug treatment for supportive treatment and measures for oxygen therapy and 6) adverse events. The clinical information was merged with selected laboratory and pharmacy information from the HIS and LIS database. All clinical, virological and laboratory data as well as adverse events were reviewed by two physicians, and all radiologic images were reviewed by two radiologists.

SARS-CoV-2 nucleic acid was detected by real-time fluorescence reverse transcriptional polymerase chain reaction (RT-PCR) using the platform of Da’an Gene Corporation, Sun Yat-sen University, Guangzhou, China. The specimens were obtained using pharyngeal swabs of patients. The nucleic acid detection of SARS-CoV-2 targeted the open reading frame 1a/b (ORF1a/b) and nucleocapsid protein (N) genes. Viral RNA was extracted with Nucleic Acid Isolation Kit on an automatic workstation Smart 32. A 200 μl sample was used for extraction following the standard protocol, and viral RNA was eluted with 60 μl elution buffer. Real-time reverse transcriptional polymerase chain reaction (RT-PCR) reagent was used following the RNA extraction. In brief, two PCR primer and probe sets, targeting ORF1ab (FAM reporter) and N (VIC reporter) genes separately, were added in the same reaction. Positive and negative controls were included for each batch. Samples were considered to be positive when either or both set(s) gave a reliable signal(s) [14].

### Outcomes

The primary outcome was the time of positive-to-negative conversion of SARS-CoV-2 nucleic acid from the initiation of treatment to day 21, with the enrollment day as the first day of treatment. The secondary outcomes included 1) the rate of positive-to-negative conversion of SARS-CoV-2 nucleic acid at day 7 of treatment; 2) the rate of positive-to-negative conversion of SARS-CoV-2 nucleic acid at day 14; 3) the rate of antipyresis (defined as axillary temperature ≤37.3°C for more than 72 hours) from the first day of treatment; 4) the rate of cough alleviation from initiation; 5) the improvement rate of chest CT at days 7 and 14; 6) the deterioration rate of clinical status from mild/moderate to severe/critical status during the study period. The severe status was defined as meeting any of the following criteria: experiencing respiratory distress, RR≥30 times/minute; oxygen saturation ≤93% in the resting state; arterial blood oxygen partial pressure (PaO2)/oxygen concentration (FiO2) ≤300 mmHg (1mmHg = 0.133kPa) [5]. The critical status was defined as meeting any of the following criteria: development of respiratory failure requiring mechanical ventilation; occurrence of shock; requirement for ICU monitoring and treatment because of complications with other organ failures [5].

Specimens from pharyngeal swabs were tested every 2 to 3 days. Negative conversion of nucleic acid was defined as negative detection of SARS-CoV-2 nucleic acid for two consecutive instances separated by more than 24 hours. Criteria of chest CT improvement included: 1) no new exudative lesions; 2) decreasing size of exudative lesions; 3) decreasing densities of lesions.

All participants were monitored for adverse events. Safety outcomes were assessed from serious adverse event reports. Any unexpected medical occurrence resulting in death, prolonged hospitalization, persistent or significant disability or incapacity, which was judged to be causally related to the study intervention, would be reported as a serious adverse event to the Institutional Review Board. Potential adverse events for the study were defined as follows (1) anaphylaxis; (2) elevation of ALT or AST to more than 2.5-fold the upper normal limit or elevation of TBIL to more than 1.5-fold the upper normal limit; (3) acute pancreatitis; and (4) diarrhea.

## QUANTIFICATION AND STATISTICAL ANALYSIS

The aim of this study is to explore the efficacy and safety of Lopinavir/ritonavir (LPV/r) monotherapy or arbidol monotherapy on the treatment of COVID-19 patients. However, COVID-19 is a new emerging disease without any data to provide a basis for calculating the sample size. In addition, the trend of the epidemic was not clear while we were designing the study. Based on the estimated number of patients admitted to the hospital at that time, we initially estimated that a maximum of 125 patients could meet the inclusion criteria, however, only 86 were ultimately recruited because few new cases developed in Guangzhou with the epidemic under control.

All statistical analyses were performed using SPSS (Statistical Package for SPSS, version 26.0). We presented continuous measurements as mean (SD) if the data were normally distributed or median (IQR) if they were not, and categorical variables as count (%). Means for continuous variables were compared using one-way ANOVA when the data were normally distributed; otherwise, the Mann-Whitney test was used. Proportions for categorical variables were compared using the χ^2^ test or Fisher’s exact tests. A two-sided α of less than 0.05 was considered statistically significant.

## KEY RESOURCE TABLE

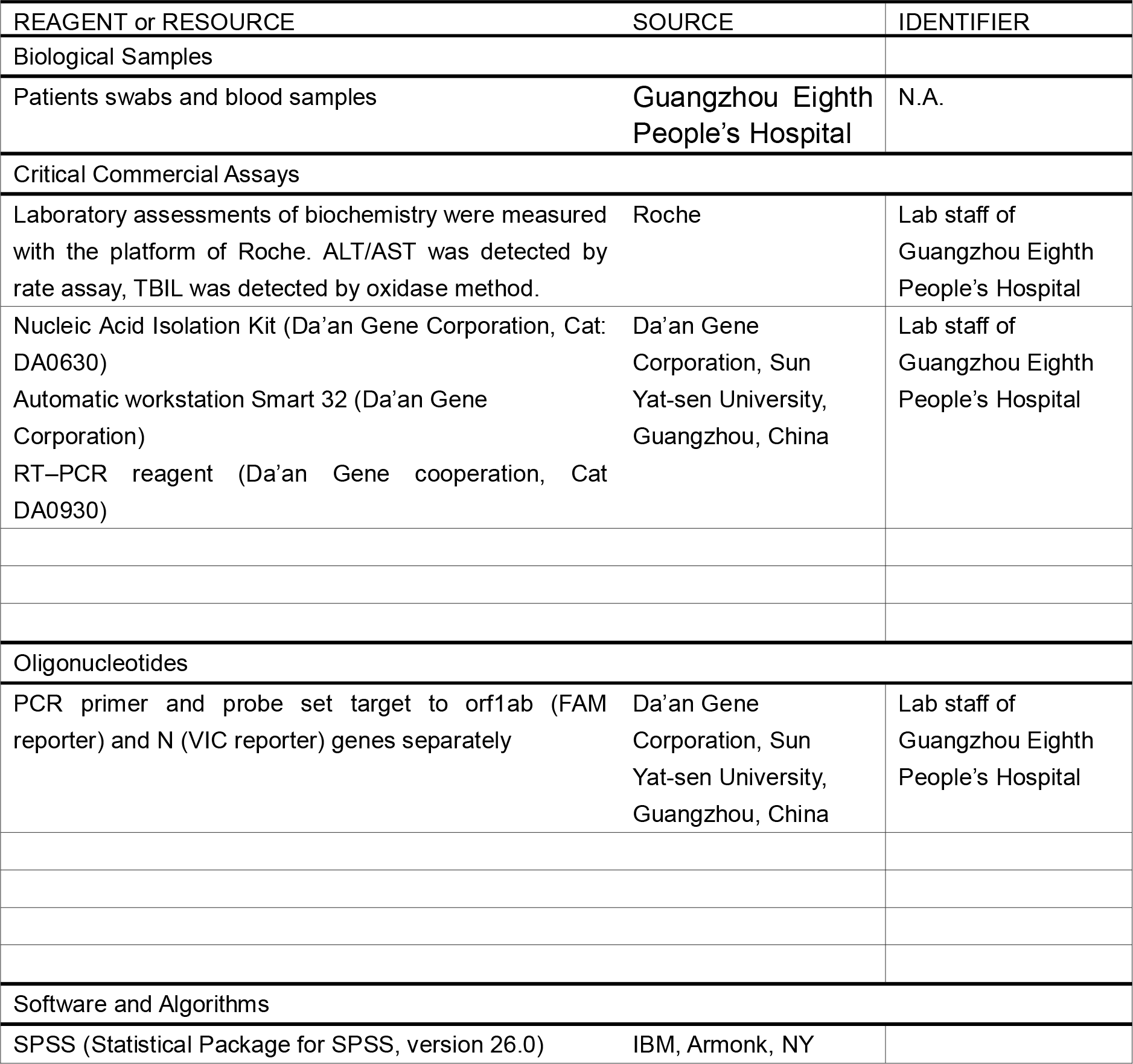
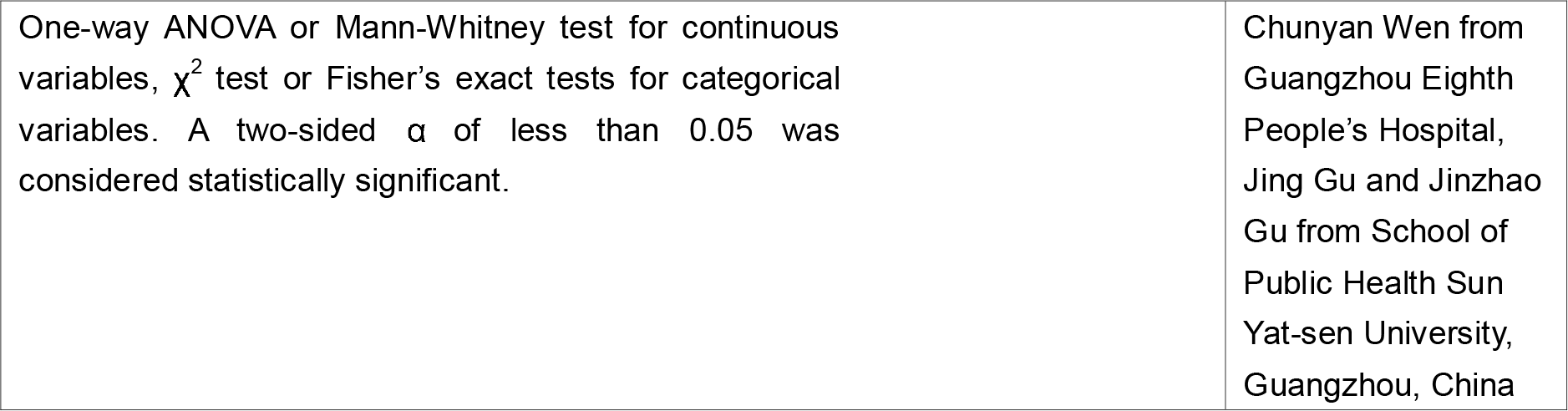

## RESULTS

### Baseline data of patients

From Feb 1 to March 28, 2020, 105 patients with COVID-19 were screened for this study, among whom 86 patients (mean age of 49.4 years [SD 14.7, range 19-79]) including 40 men and 46 women were successful enrolled (figure 1). Patients were randomly assigned to receive LPV/r (n=34, arbidol (n=35), or control (n=17). All patients were followed up for 21 days.

**Figure 1.**
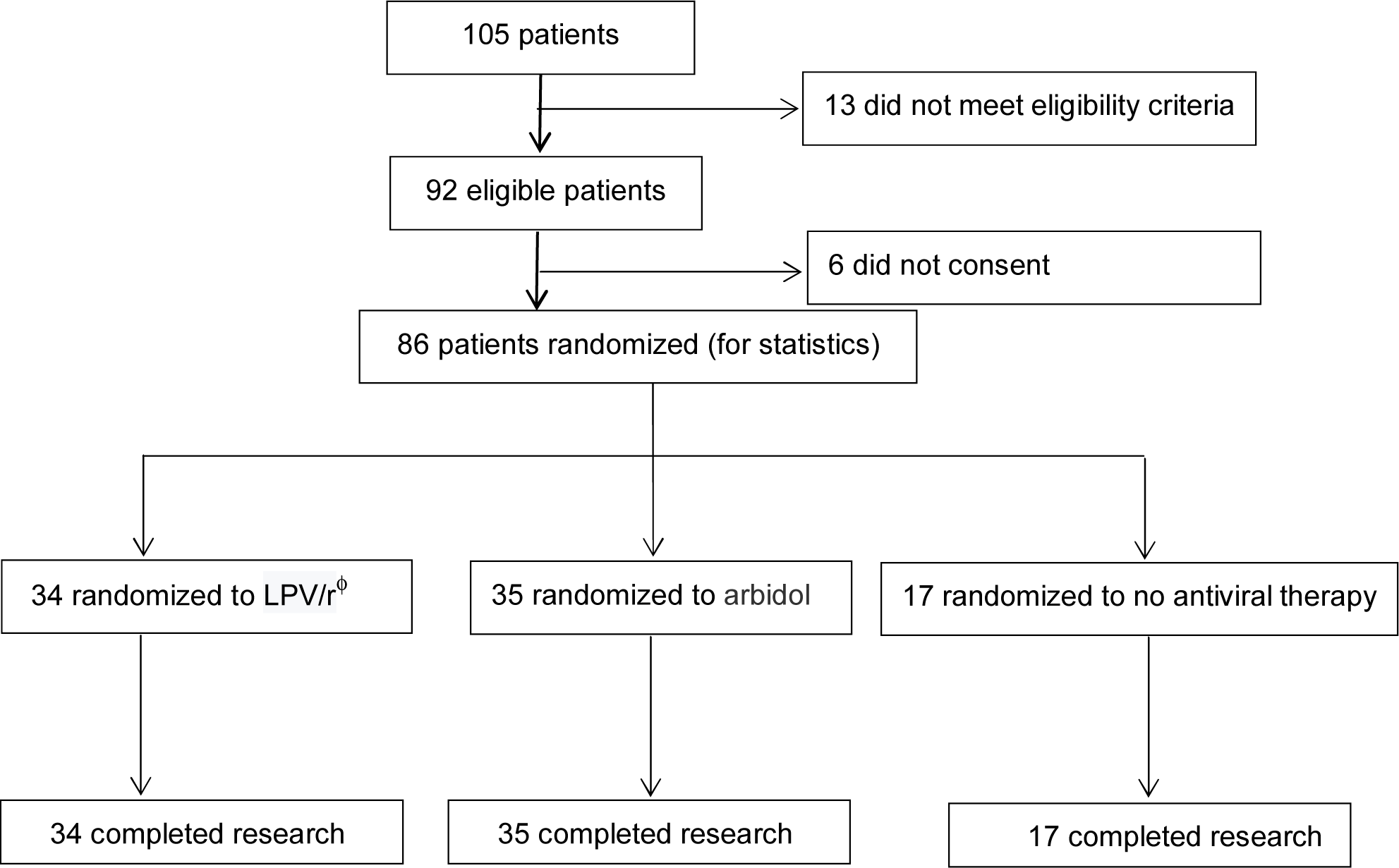
Trial profile ^θ^ SAE: Severe Adverse Event ^ϕ^ LPV/r: Lopinavir/ritonavir

None of the enrolled patients had chronic lung disease, chronic kidney disease, autoimmune disease or immunodeficiency disease. Twenty-seven (79.4%) patients in the LPV/r group, 22 (62.9%) in the arbidol group and 9 (52.9%) in the control group experienced fever. Twenty-one (61.8%) patients in the LPV/r group, 25 (71.4%) in the arbidol group and 9 (52.9%) in the control group developed cough. There were no significant differences in baseline demographic data, common clinical manifestations or pneumonia incidence seen on chest CT imaging between the three groups (*P*>0.05). The baseline characteristics of the 86 patients in three groups are shown in table 1. None of the patients of complained dyspnea, diarrhea, palpitation or headache on admission. The laboratory parameters of alanine aminotransferase (ALT), aspartate aminotransferase (AST), total bilirubin (TBIL) and creatinine were normal when patients started the antiviral treatment. Other laboratory parameters including white blood cell count, lymphocyte count, neutrophil count, C-reactive protein level and procalcitonin level did not show significant differences between the three groups (*P* >0.05).

**Table 1:** Baseline characteristics of the three treatment groups (intention-to-treat population)

| Characteristics | LPV/r<br>( n=34 ) | Arbidol<br>( n=35 ) | Control<br>( n=17 ) | P value |
| --- | --- | --- | --- | --- |
| Gender (n, %) |  |  |  | 0.831 |
| Male | 17(50.0%) | 16(45.7%) | 7(41.2%) |  |
| Female | 17(50.0%) | 19(54.3%) | 10(58.8%) |  |
| Age, in years (mean, SD, range) | 50.7(15.4,19-79) | 50.5(14.6,20-74) | 44.3(27-62) | 0.288 |
| Time from onset to treatment, in days (median, IQR) | 3.5(2,6) | 6(2,8) | 5(2,8) | 0.371 |
| Underlying chronic diseases<sup>¶</sup> (n, %) | 7(20.6%) | 5(14.3%) | 6(35.3%) | 0.236 |
| Evidence of pneumonia based on chest CT imaging (n, %) | 28(82.4%) | 33(94.3%) | 14(82.4%) | 0.234 |
| Clinical status (n, %) |  |  |  | 0.234 |
| mild | 6(17.6%) | 2(5.7%) | 3(17.6%) |  |
| moderate | 28(82.4%) | 33(94.3%) | 14(82.4%) |  |
| White blood cell count,10<sup>9</sup>/L |  |  |  | 0.671 |
| <4 (n, %) | 8(23.5%) | 11(31.4%) | 3(17.6%) |  |
| 4-10 (n, %) | 25(73.5%) | 24(68.6%) | 14(82.4%) |  |
| >10 (n, %) | 1(2.9%) | 0 |  |  |
| Lymphocyte count,10<sup>9</sup>/L |  |  |  | 0.974 |
| <1.1 (n, %) | 9(26.5%) | 9(25.7%) | 4(23.5%) |  |
| 1.1-3.2 (n, %) | 25(73.5%) | 26(74.3%) | 13(76.5%) |  |
| Neutrophil<br>count, 10<sup>9</sup>/L |  |  |  | 0.981 |
| <1.8 (n, %) | 5(14.7%) | 6(17.1%) | 2(11.8%) |  |
| 1.8-6.3 (n, %) | 26(76.5%) | 27(77.1%) | 14(82.4%) |  |
| >6.3(n, %) | 3(8.8%) | 2(5.7%) | 1(5.9%) |  |
| C-reactive<br>protein, mg/L |  |  |  | 0.364 |
| <10 (n, %) | 17(50.0%) | 23(65.7%) | 11(64.7%) |  |
| >10(n, %) | 17(50.0%) | 12(34.3%) | 6(35.3%) |  |
| Procalcitonin, ng/mL |  |  |  | 0.308 |
| <0.05 (n, %) | 18(52.9%) | 24(68.6%) | 12(70.6%) |  |
| >0.05(n, %) | 16(47.1%) | 11(31.4%) | 5(29.4%) |  |
| Use gamma globulin<br>(%) | 3/34(8.8%) | 2/35(5.7%) | 1/7(5.9%) | 0.865 |
| Use of<br>glucocorticoids (%) | 7/34(20.6%) | 2/35(5.7%) | 3/17(17.6%) | 0.153 |
| Oxygen therapy (%) |  |  |  | 0.677 |
| None | 12/34(35.3%) | 13/35(37.1%) | 7/17(41.2%) |  |
| Low flow oxygen<br>supply | 15/34(44.1%) | 19/35(54.3%) | 8/17(47.0%) |  |
| High flow oxygen<br>supply | 7/34(20.6%) | 3/35(8.6%) | 2/17(11.8%) |  |

† list some chronic diseases, including one patient with diabetes, 4 hypertension and 2 with both in the LPV/r group; one patient with Coronary Heart Disease, 3 hypertension and 1 chronic liver disease in the arbidol group; and one patient with Coronary Heart Disease, 2 hypertension, 2 chronic liver disease and 1 with both diabetes and chronic liver disease in the control group.

During the study period, six patients used gamma globulin (10 g, once a day, for 2-3 days),12 used glucocorticoids (methylprednisolone 40 mg, once a day, for 3-5 days) and 54 received oxygen supply therapy. The usage percentages of the above supportive treatment did not show statistical differences between the three groups (*P*=0.865, *P*=0.153, *P*=0.677 respectively).

### Efficacy outcomes

The mean time for positive-to-negative conversion of SARS-CoV-2 nucleic acid was 9.0 days (SD 5.0) in the LPV/r group, 9.1 (SD 4.4) in the arbidol group and 9.3 (SD 5.2) in the control group, with no statistical difference between them (*P*=0.981) (table 2 and figure 2). After 7 days of treatment, the positive-to-negative conversion rates of SARS-CoV-2 nucleic acid in pharyngeal swab in the LPV/r group, the arbidol group and the control group were 35.3%(12/34), 37.1% (13/35) and 41.2% (7/17) respectively and did not present statistical difference among the three groups (*P*=0.966) (table 2). After 14 days of treatment, the positive-to-negative conversion of SARS-CoV-2 nucleic acid was 85.3% (29/34), 91.4% (32/35) and 76.5% (13/17) respectively in the LPV/r group, the arbidol group and the control group, without significant statistical difference between them (*P*= 0.352) (table 2). Over the 21-day follow-up, the cumulative incidence of positive-to-negative conversion of SARS-CoV-2 nucleic acid in pharyngeal swabs did not show statistical difference between the three groups (figure 3).

**Table 2:**
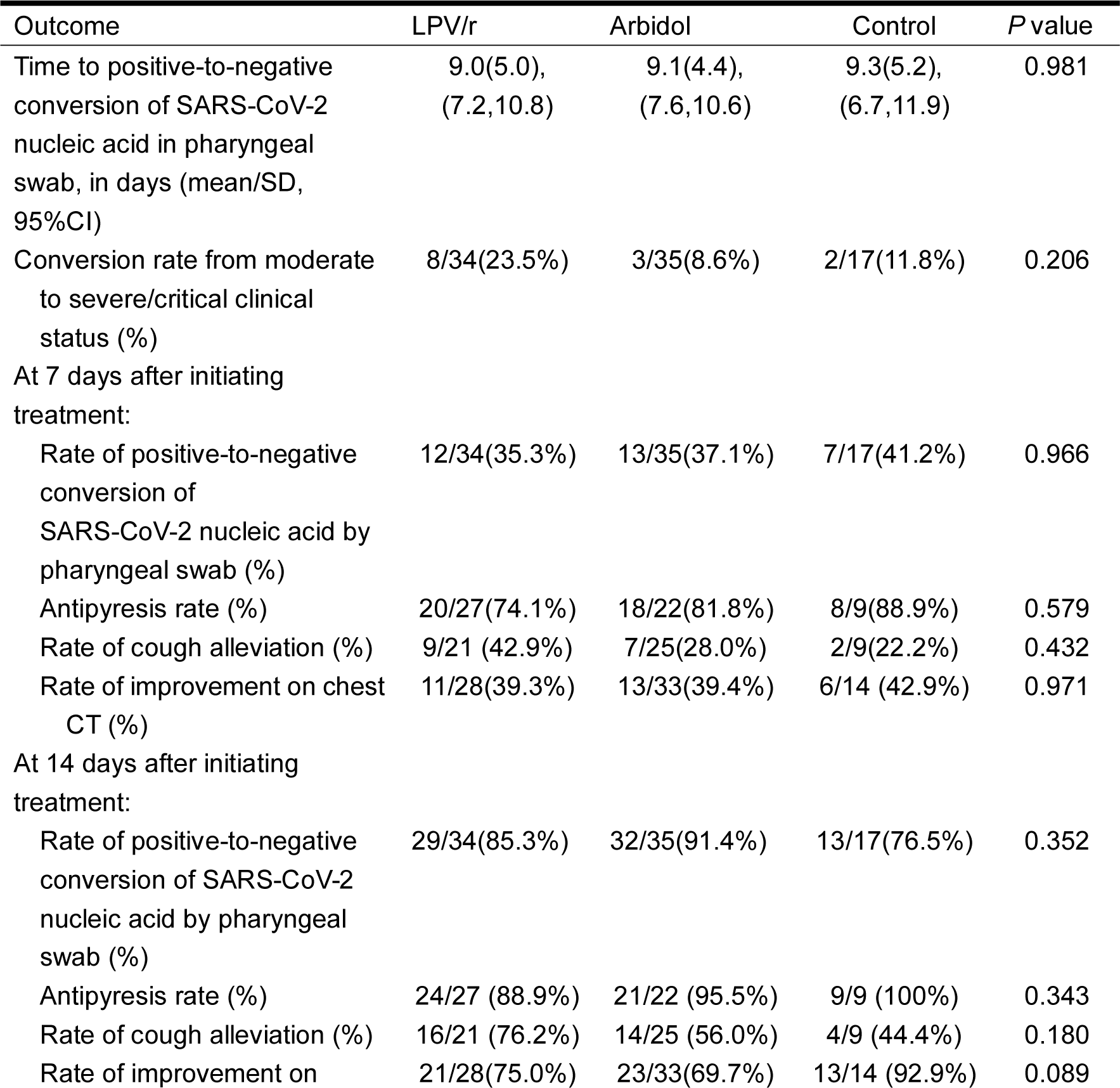
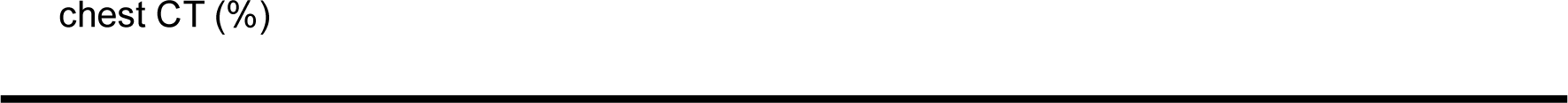
Outcomes of the three groups (intention-to-treat population)

**Figure 2.**
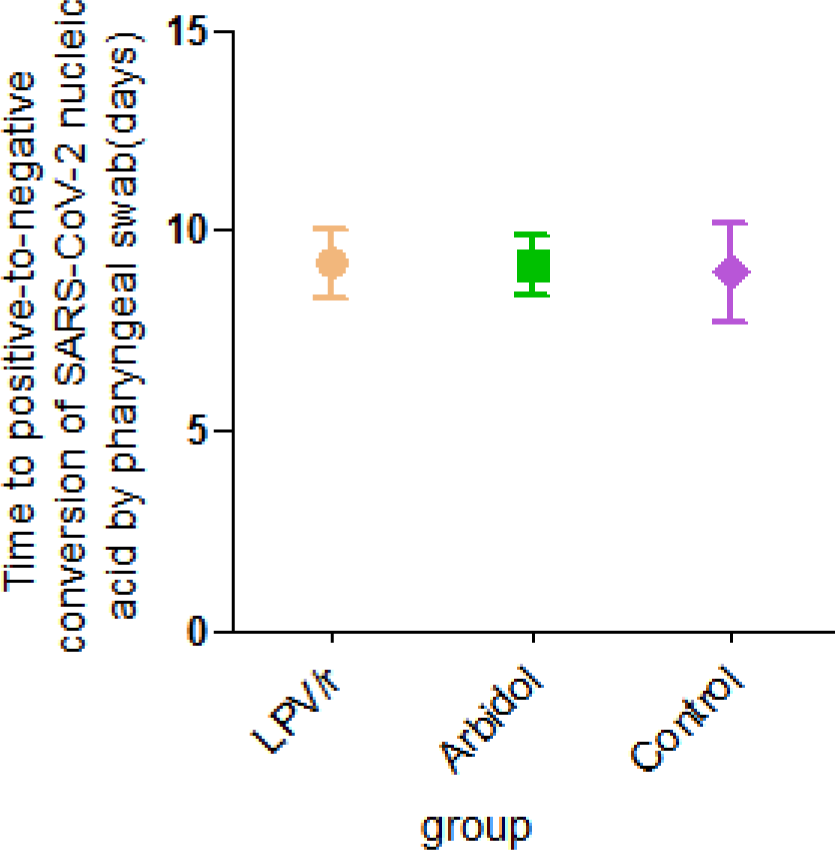
Time to positive-to-negative conversion of SARS-CoV-2 nucleic acid by pharyngeal swab in each of the treatment three groups during the 21-day follow-up period LPV/r: lopinavir/ritonavir SARS-CoV-2: severe acute respiratory syndrome coronavirus 2

**Figure 3.**
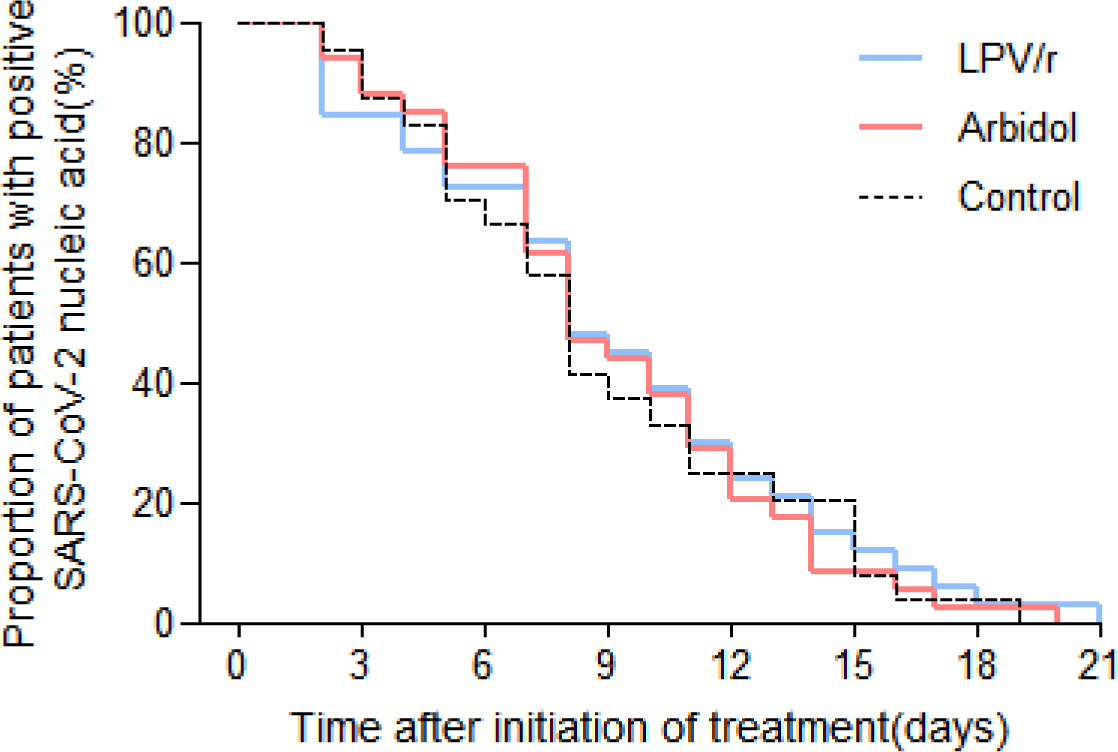
Proportion of patients in each of the three treatment groups with positive SARS-CoV-2 nucleic acid by pharyngeal swab during the 21-day follow-up period LPV/r: lopinavir/ritonavir SARS-CoV-2: severe acute respiratory syndrome coronavirus 2

With respect to other secondary outcomes, the rate of antipyresis, rate of cough resolution, and rate of improvement on chest CT imaging at day 7 and 14 did not show any statistical difference between the three groups (*P* > 0.05). At day 7, eight (23.5%) patients in the LPV/r group, 3 (8.6%) in the arbidol group and 2(11.8%) in the control group deteriorated from mild/moderate clinical status to severe/critical clinical status, without statistical difference (*P* =0.206) (table 2).

In order to rule out the influence of the time from onset to treatment on the clinical status, we compared the time from onset to treatment in patients who deteriorated to severe/critical clinical status [5 (IQR 2, 8) days] with those who did not [4 (IQR 2, 7) days], and did not find any significant difference between them (*P* =0.619).

### Safety outcomes

During the follow-up period, 12 (35.3%) patients in the LPV/r group experienced adverse events including diarrhea (9/34, 26.5%), loss of appetite (5/34, 14.7%) and elevation of ALT over 2.5-fold above the normal limit (1/21, 4.8%). In addition, 5 (14.3%) patients in the arbidol group experienced adverse events including diarrhea (3/35, 8.6%) and nausea (2/34, 5.9%). No apparent adverse events occurred in the control group. Notably, one serious adverse event occurred in a 79-year-old man with underlying diseases including diabetes and hypertension in the LPV/r group, characterized by severe diarrhea on day 3. The patient withdrew from this study and tested positive for SARS-CoV-2 nucleic acid lasting over 14 days of the follow-up period. This patient progressed to critical condition and received extracorporeal membrane oxygenation (ECMO). Fortunately, he recovered and stopped needing ECMO by the observation endpoint of this study.

### Summary of cases with severe/critical clinical status

A total of 13 (15.1%) patients (8 men, 5 women) progressed to severe/critical clinical status (containing 11 severe cases and 2 critical cases) during the study period including 8 receiving LPV/r, 3 receiving arbidol and 2 control. The two critical cases belonged to LPV/r group.. The mean age of these 13 patients was 60.1 years [SD 13.8, range 37-79],. Ten (76.9%) patients came from Hubei province and 3 (23.1%) were local residents of Guangzhou. Two (15.4%) patients suffered from diabetes mellitus, 5 (38.5%) from hypertension and 1 (7.7%) from chronic liver disease. All patients complained of fever and 9 of them (69.2%) complained of cough, but none experienced diarrhea at the beginning of treatment. The SaO2 at rest was ≤93% in 4 (30.8%) patients and PaO2/FiO2 ratio was ≤300 in 5 (38.5%) patients. Among these patients, 2(15.4%) required mechanical ventilation due to respiratory failure. Five (38.5%) and 9 (69.2%) cases achieved positive-to-negative conversion of SARS-CoV-2 nucleic acid at days 7 and 14 respectively. At days 7 and 14 of follow-up, 6 (46.2%) and 10 (76.9%) patients had improvements in chest CT imaging. At the follow-up endpoint of day 21, 12 patients had been discharged from hospital and only one case was still hospitalized. No deaths occurred.

## Discussion

Several clinical studies have reported the treatment of a large number of COVID-19 patients with antiviral and antibiotic therapy [15-17]. However, no specific medication has proven effective for suppressing or eliminating SARS-CoV-2 infection or for reducing complications and mortality. There are several ongoing clinical drug trials registered in the Chinese clinical trial registry [15,18]. Although the epidemic within China is now largely under control, epidemics in other countries are becoming increasingly severe [2]. Therefore, it is extremely important to find specific anti SARS-CoV-2 drugs and learn from the experience of Chinese health providers.

Our study was designed to be an empirical exploration intended to recruit 125 adult patients hospitalized with mild/moderate COVID-19; however, only 86 patients were involved in this study for the reasons previously mentioned. Through randomization, 34 patients were assigned to receive LPV/r, 35 to arbidol, and 17 to no antiviral medication as control. The results showed that LPV/r and arbidol did not shorten the time of positive-to-negative conversion of COVID-19 nucleic acid in respiratory specimens (9.0 vs. 9.1 vs. 9.3 days), nor did they improve the symptoms of COVID-19 or pneumonia on lung CT imaging at 7 days and 14 days. Moreover, more patients treated with LPV/r progressed from mild/moderate to severe/critical status than patients from the other two groups

One reason behind the failure of LPV/r and arbidol to improve patient outcome could be that a higher dose is needed to successfully suppress SARS-CoV-2 in patients to achieve an effect comparable to to in vitro cytotoxicity tests [8, 13]. However, this would be difficult to achieve clinically given the side effects caused by both drugs. In particular, it should be noted that patients treated with LPV/r had more gastrointestinal symptoms, which might affect the patient’s recovery. Based on the drug instruction and previous experience in treating HIV-infected patients, the adverse reactions of the short-term use of LPV/r mainly include diarrhea, abnormal stools, abdominal pain, nausea, vomiting, and asthenia [6]. Since the above side effects may aggravate the disease, LPV/r treatment should be cautiously considered after weighing the risks and benefits.

The results in our study are consistent with findings from a recent clinical trial of LPV/r in adults hospitalized with severe COVID-19 conducted in Wuhan, which recruited 199 hospitalized adult patients with severe COVID-19 and concluded that no benefit was observed with LPV/r treatment beyond standard care [19]. In addition, another retrospective clinical research study conducted in Shanghai observed 134 patients with COVID-19 and did not find any effects of LPV/r and arbidol on relieving symptoms or accelerating virus clearance after treatment for 5 days [20]. Despite the small sample size, our study also suggests that monotherapy of LPV/r or arbidol might not improve the clinical outcome in treating with mild/moderate COVID-19.

During the study period, a total of 13 (15.1%) patients progressed to severe/critical clinical status including 8 receiving LPV/r, 3 receiving arbidol and 2 in the control group, which indicates that disease condition could still worsen even after hospitalization, and thus urgently demands rigorous observation of illness and care. Fortunately, by the endpoint of this study twelve patients had been discharged from hospital after recovery, and only one patient remained hospitalized, albeit with significant clinical improvement. This gives us confidence that even in the absence of specific antiviral drugs, the vast majority of COVID-19 patients in severe/critical clinical status can still recover after comprehensive treatment.

### Limitations of study

Our study is not without its limitations. First, we recognize that our sample size was small. Second, the study did not enroll severely or critically ill patients, or patients at increased risk of poor outcome with many comorbidities and was conducted in only one center. Third, the study was not completely blinded, possibly influencing the outcome to some extent. We will continue to follow these patients to evaluate their long-term prognosis. Nevertheless, as a prospective randomized, controlled trial, this study could still provide meaningful suggestions for proper application of LPV/r and arbidol in the treatment of COVID-19.

In conclusion, our study found that LPV/r or arbidol monotherapy presents little benefit for improving the clinical outcome of hospitalized patients with mild/moderate COVID-19 beyond symptomatic and supportive care, causing instead more adverse events. Further work is needed to confirm these results.

## Data Availability

All data included in this study are available upon request by contact with the corresponding author.

## Contributors

Linghua Li, Xilong Deng and Yueping Li conceived the study and designed the protocol. Weiping Cai and Fuchun provided oversight. Chunyan Wen contributed to statistical analysis and interpretation of data. Linghua Li and Weiyin Lin drafted the manuscript. Feng Li and Fengyu Hu conducted of nuclear acid RT-PCR. Zhiwei Xie and Yujuan Guan reviewed the data independently. Jinxin Liu and Lieguang Zhang reviewed all radiologic images independently. Xiaoneng Mo, Jian Wang, Yaping Wang, Ping Peng, Xudan Chen, Wenxin Hong and Guangming Xiao contributed to conducting the study and collecting data.

## Acknowledgments

This study was supported by Chinese 13th Five-Year National Science and technology major projec(t 2018ZX10302103-002, 2017ZX10202102-003-004), and Infectious Disease Specialty of Guangzhou High-level Clinical Key Specialty (2019-2021). We thank all patients who participated in this study and all staff of Guangzhou Eighth People’s Hospital for their clinical care given to patients and facilitating access to the relevant medical records. We also thank Jing Gu and Jinzhao Gu from School of Public Health Sun Yat-sen University, Guangzhou, China for statistical assistance, and Nancy Yang from University of Minnesota Twin Cities for English language assistance.

